# The Mini Linguistic State Examination (MLSE): a brief but accurate assessment tool for classifying Primary Progressive Aphasias

**DOI:** 10.1101/2020.06.02.20119974

**Authors:** Nikil Patel, Katie A. Peterson, Ruth Ingram, Ian Storey, Stefano F. Cappa, Eleonora Catricala, Karalyn E. Patterson, Matthew A. Lambon Ralph, James B. Rowe, Peter Garrard

**Affiliations:** Molecular and Clinical Sciences Research Institute, St George’s, University of London, SW17 0RE; Department of Clinical Neurosciences and Cambridge University Hospitals NHS Trust, University of Cambridge, CB2 0SP; Division of Neuroscience and Experimental Psychology, University of Manchester, M13 9PL; University Institute for Advanced Studies IUSS, Pavia, Italy; IRCCS Mondino Foundation, Pavia, Italy; MRC Cognition and Brain Sciences Unit, University of Cambridge, CB2 7EF

**Keywords:** Primary progressive aphasia, semantic dementia, progressive nonfluent aphasia, logopenic progressive aphasia, Alzheimer’s disease, frontotemporal dementia

## Abstract

**Background:** This paper introduces a new clinical test, the Mini Linguistic State Examination (MLSE), as a short assessment for screening and classification of the different manifestations of primary progressive aphasia (PPA). Differentiation and monitoring of PPA variants are vital for management, planning and development of new treatments. The MLSE is designed to improve the uniformity of testing, screening for recruitment to clinical trials, and consistency of research results. It is a brief but effective test which can be adapted to the world’s major languages.

**Methods:** Fifty-four patients and 30 age-, sex- and education-matched controls completed testing with the MLSE and components of the Boston Diagnostic Aphasia Examination in addition to their standard clinical diagnostic assessment. The MLSE includes five domains (motor speech, phonology, semantics, syntax and working memory) that were compared across groups. A random forest classification was used to learn the relationship between these five domains and assess the power of the diagnostic accuracy for predicting PPA subtypes. The final machine learning model was used to create a decision tree to guide the optimal manual classification of patients.

**Results:** On average, the test took less than 20 minutes to administer. Significant group differences were found across all five domains, in terms of the distributions of error-types. These differences mirror the well-known language profiles for the three main PPA variants, which typically require an extended neuropsychology and speech pathology assessment. The random forest prediction model had an overall classification accuracy of 96% (92% for logopenic variant PPA, 93% for semantic variant PPA and 98% for non-fluent variant PPA). The derived decision tree for manual classification produced correct classification of 91% of participants whose data were not included in the training set.

**Conclusions:** The MLSE is a new short cognitive test, with a scoring system that is easy to learn and apply. It is accurate for classifying PPA syndromes, and has potential to screen and monitor language deficits that occur in other focal and neurodegenerative brain disorders associated with language impairment. With increasing importance of language assessment in clinical research, the MLSE’s linguistic assessment tool enables the essential profiling of language deficits in a wide clinical community.

## Introduction

Alzheimer’s disease (AD) and frontotemporal dementia (FTD) can present with difficulty in language production and/or comprehension as the dominant, or even sole, clinical feature. Collectively, such language presentations of neurodegenerative conditions are referred to as ‘primary progressive aphasia’ (PPA) (Mesulam, 1982). The World Federation of Neurology working group defined three distinct subtypes of the phenomenon: progressive non-fluent variant (nfvPPA), characterised by effortful and/or agrammatic language production; semantic variant (svPPA), presenting with anomia and problems with single word comprehension; and logopenic variant (lvPPA), in which word retrieval and sentence repetition deficits are prominent (Gorno-Tempini *et al*., 2011).

The core features distinguishing svPPA, nfvPPA and lvPPA can be reliably detected and quantified using validated test batteries such as the Boston Diagnostic Aphasia Examination (BDAE) or the Western Aphasia Battery (WAB) (Flamand-Roze *et al*., 2011; Kertesz A, 1982). Administration and interpretation of such procedures is, however, time-consuming and depends on specialist expertise that is not widely accessible. In practice, clinical classification is therefore more often based on an informal assessment, which will inevitably lead to inconsistencies and still requires specialist skills. Such inconsistency and dependence on centralised specialists will impede the scalability of language screening, aphasia diagnostics, and the development of disease modifying therapies. There therefore is a pressing need for harmonisation of scalable, quantifiable clinical description and diagnosis of aphasias.

One such bedside language battery that has been proposed is the Italian ‘SAND’ (Screening for Aphasia in NeuroDegeneration) (Catricala *et al*., 2017). Several aphasia scales in English(e.g. PARIS, PASS and PALS) lead to standardised estimates of clinical severity, but are not designed to distinguish the three main subtypes of PPA. Similarly, brief aphasia batteries such as the Comprehensive Aphasia Test (Swinburn *et al*., 2004), the Aphasia Rapid Test (Azuar *et al*., 2013), the Language Screening Test (Flamand-Roze *et al*., 2011), and the Frenchay Aphasia Screening Test (Enderby and Crow, 1996), were designed primarily to assess aphasia after stroke, and are not explicitly oriented to probe the distinguishing features of the PPA syndromes. An alternative approach would be a formal analysis of connected speech samples using the approaches described by Ash (2013) or Fraser (2014) and their coworkers, but unless fully automated, the procedure would be too onerous for clinical practice and operator dependent in quantification. Savage et al., (2013) proposed that four language tasks – picture naming, single-word comprehension, semantic association and single word repetition – could distinguish PPA syndromes with reasonable reliability. The resulting “Sydney Language Battery” classified 90% of svPPA cases, 84% of nfvPPAs, and 73% of the lvPPAs in accordance with their clinical diagnoses, with 80% of PPA correctly classified overall (Savage *et al*., 2013). However, in the absence of tests of grammatical processing, the battery would not reliably distinguish newly diagnosed nfvPPA, while the absence of sentence repetition may lead to the misclassification of lvPPA.

A comprehensive assessment of language in patients with PPA would include measures of the motor aspects of speech production, knowledge of the meaning and phonological properties of words and the syntactic structure of sentences, in addition to confrontation naming and semantic association. A test of auditory verbal working memory is also required, given its crucial role in many verbal tasks, such as repetition and sentence comprehension. We therefore developed and validated a new brief clinical instrument that would allow clinicians to profile multiple aspects of linguistic competence in a consistent, quantitative and reproducible way.

In designing the Mini Linguistic State Examination (MLSE) we aimed to ensure that it would be: i) usable after minimal training, with no specialised knowledge of aphasiology; ii) brief enough to form part of an outpatient or ward-based clinical assessment of suspected or established cases; iii) sensitive to the differing patterns of linguistic deficit characterising the three archetypal syndromes of PPA; and iv) able to capture atypical symptom clusters. Finally, and in a departure from conventional clinical scoring methods based on response accuracy, we proposed that recording the rates at which different types of error were made by a participant would yield a high level of discrimination.

In this paper we present the MLSE and report the profiles obtained in a cohort of patients with predominantly mild PPA, recruited through specialist cognitive neurology services at three centres. We report statistics relating to the validity, reproducibility, accuracy and ease of administration of the MLSE, and the output of a machine learning derived decision tree to classify the PPA subtypes using data obtained from administering the test.

## Methods

### Participants

A total of 61 patients with PPA (25 lvPPA, 20 nfvPPA, 16 svPPA) were recruited through specialist cognitive neurology clinics at St George’s Hospital, London (*n* = 26), Addenbrooke’s Hospital, Cambridge (*n* = 27), and Manchester Royal Infirmary and its associated clinical providers (*n* = 8). Diagnosis was based on the WFN working group criteria (Gorno-Tempini *et al*., 2011), including brain imaging, neuropsychological assessment and clinical review by multidisciplinary teams including aphasiology expertise. Three patients declared a native language other than English but all were highly fluent, had been communicating in English since childhood, and predominantly or exclusively used English in day-to-day life. Three patients and four controls subjects were left-handed. Seven patients were excluded due to the advanced stage of their condition (4 x lvPPA, 3 x nfvPPA) leaving 54 PPA patients in the final analysis.

Thirty healthy volunteers were recruited through the National Institute for Health Research “Join Dementia Research” registers in London and Cambridge, and invitations to patients’ relatives. Controls had no history of significant neurological, psychological, speech and language, or learning deficits. All were native speakers of English with normal or corrected-to-normal hearing and vision.

Written informed consent was provided by all participants. The study protocol was reviewed and approved by the London (Chelsea) Research Ethics Committee [REC reference: 16/LO/1735]. The study was sponsored by St George’s, University of London, the University of Cambridge and the University of Manchester.

### Experimental design

Participants underwent baseline assessments including the Addenbrooke’s Cognitive examination (ACE-III) (Hsieh *et al*., 2013) and the short form of the BDAE (Goodglass H., 2001). If a participant had completed the ACE-III within a month prior to performing the MLSE, the ACE-III version B was administered. The assessment session was video and/or audio recorded for offline scoring and analysis.

### The MLSE

The MLSE consists of eleven subtests, each of which makes a different combination of demands on the components of language competence that are affected by PPA (Gorno-Tempini *et al*., 2011). As there is no individual test of language production or comprehension that is selectively sensitive to any of these components in isolation, the MLSE captures the nature of a patient’s language impairment on the basis of the number and nature of error responses.

Five error-types are considered. These reflect dysfunction of: i) the motoric aspects of speech; ii) semantic knowledge; iii) knowledge of phonology; iv) knowledge of syntax, and v) auditory-verbal working memory. The method generates a profile score that reflects performance within five domains of linguistic competence, as well as an overall score reflecting the severity of the language disorder.

General definitions of the five error types are provided in Table 1. Additionally, because the circumstances under which errors occur differ depending on the task being performed (e.g., between written and spoken tasks, or between those that require verbal vs. non-verbal responses), definitions relevant to each of the eleven subtests are also specified, and illustrated with examples, in the MLSE administration and scoring guide (see supplemental material A).

**Table 1.** General definitions of the five types of errors that are recorded during administration of the MLSE.

| Error type | Definition | Notes |
| --- | --- | --- |
| **Motor speech** | A response that is slurred, stuttered or contorted, and which the examiner would find difficult to repeat or transcribe. | Motor speech errors arise only during tasks requiring speech production. A motor speech error should be noted and scored, even when self-corrected. The errors are not confined to speech dyspraxia. |
| **Phonological** | A response that contains incorrect but word-like components, and which could easily be repeated or written down. | Phonological errors arise only during tasks requiring speech production. Any phonological error should be noted and scored, even when self-corrected. |
| **Semantic** | A semantic error is noted when a participant's response suggests a deficit at the level of conceptual knowledge and/or word meaning. | Semantic errors can arise during both production (e.g., naming) and comprehension (e.g., picture association) tasks. Context-specific guidance is provided for each subtask. |
| **Syntactic** | A syntactic error occurs when a participant demonstrates difficulty understanding or producing grammatically correct sentences. | Context-specific guidance is provided for each subtask. |
| **Working memory** | Working memory errors are recorded when a participant is unable to repeat sentences correctly. The shorter the incorrectly repeated sentence, the higher the error score. | Working memory errors are scored only during the sentence repetition task. |

A guide featuring detailed administration and scoring instructions can be downloaded from the MLSE website (http://www.mlsexam.com) where an interactive training module is also available.

### Scoring the MLSE

A participant’s profile was determined by subtracting the number of errors of each type from the number of opportunities to make that type of error. If a participant made no errors, the test would yield a profile score of 30/30 for motor speech, 30/30 for phonology; 20/20 for semantics, 10/10 for syntax, 10/10 for working memory, and an overall score of 100/100. Multiple error types can be associated with a single response: for instance in the naming task, if a participant were to produce a semantic substitution that contained a phonological error, both a semantic and a phonological error would be recorded (see supplemental material A).

Some patients with advanced PPA were unable to make any response, even after being encouraged by the tester to make an attempt. When this occurs, the test item is associated with a ‘no-response’ error, which is equivalent to the sum of all possible domain error scores for that item. The seven PPA patients excluded from the analysis were those whose scores included ‘no-response’ errors. Example scoring of the ‘no-response’ errors can be found in supplemental material A under Figure 1.

**Figure 1.**
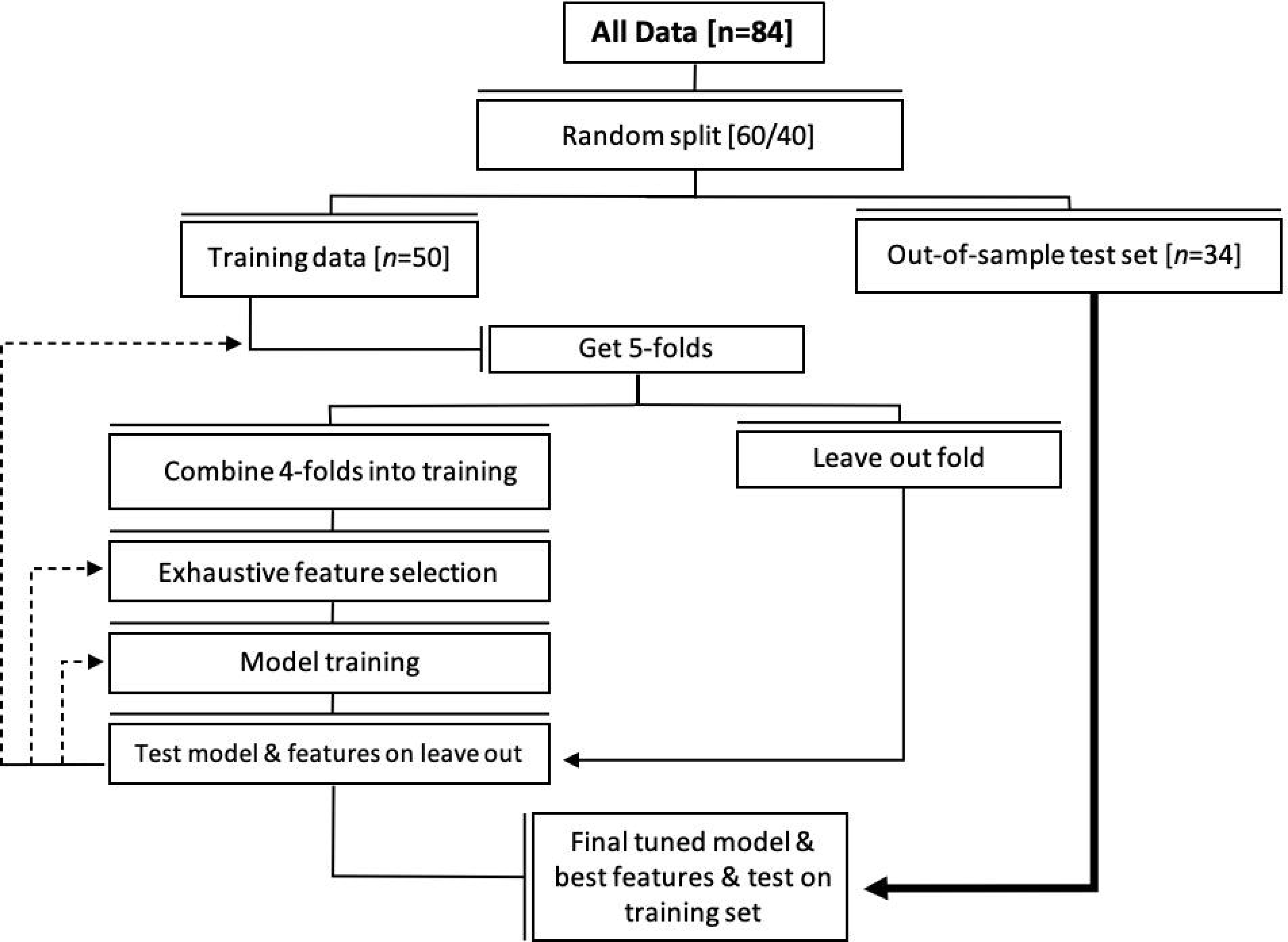
Flow diagram of random forest training and feature selection for PPA subtype classification.

Testing was performed in a quiet environment, with all sessions video and/or audio recorded to enable offline scoring and between-rater agreement measures. Recordings of thirty patient evaluations were used to perform independent parallel evaluations by three different raters (one from each site) blinded to the syndromic diagnosis.

### Statistical analysis

Data were analysed using IBM SPSS (version 25.0). Convergent validity was measured using Cronbach’s alpha (McDowell, 1996) and through correlation of standardised scores obtained in subtasks of the MLSE with relevant subsections of established measures (BDAE and ACE-III/R). Specifically, correlations between the following pairs of tests (components of the BDAE and MLSE respectively) were conducted: repetition of single words and the repetition component of the repeat and point task; auditory comprehension and the pointing component of the repeat and point task; repetition of sentences and the sentence repetition task; the Boston naming test and the naming task; oral reading and the reading task; working memory components of the ACE-III/R with the sentence repetition task. The working memory components of the ACE was the sum of the scores achieved on i) word-list repetition (but not recall), ii) sentence repetition, and iii) name and address repetition (but not recall).

Inter-rater reliability was obtained using a random intraclass correlation (ICC) model based on absolute agreement. Demographic characteristics and all test-derived scores were compared across groups using Welch’s ANOVA due to unequal variances and sample size per group (giving the asymptotically F distributed score), and *post hoc* pairwise comparisons subject to Bonferroni correction. Socio-demographic variables were compared using parametric or non-parametric tests depending on Levene’s test for equality of variance. Receiver operating characteristic (ROC) curves were plotted to assess the differential diagnostic efficiency of different features. Discriminant function analysis was conducted to demonstrate classification accuracy of the three PPA subtypes.

### Machine-learning classification

A random forest classifier was trained and tested using MATLAB (2019a, version 25.0). The random forest classification method has been applied extensively to medical data (Sarica *et al*., 2017) because of its accuracy, robustness to noisy datasets and relative immunity to overfitting (Breiman, 2001). The full sample was split randomly (weighted by the numbers in each diagnostic group) into a training (60%, n = 50) and out-of-sample test set (40%, n = 34). The training test was used for training the model using five-fold leave-one-out cross-validation. The trained model was then evaluated against the out-of-sample data,Figure 1.

The number of decision trees in the random forest was one hundred (*n* = 100). This was determined through a grid search in which a range of forests are grown containing *n* trees, where *n* begins at 10 and increments to a maximum of 1000. The number of predictors to sample was set equal to the square root of total number of features, as suggested by Liaw et al, (2002). Sensitivity, specificity, F1-score, precision, recall and balanced classification accuracy were used as evaluation metrics of average fold performance for each experiment, as well as final model testing, after manual selection of domain combinations with high balanced accuracy. The final tree structure is identified by testing each decision tree within the forest and calculating the average and variance between class accuracies of the out of sample testing data. The final model was also used to create a clinical decision tree to guide the manual classification of new test data.

### Data availability statement

Anonymised data are available on reasonable request for academic purposes.

## Results

### Participants

Group characteristics are displayed in Table 2. Age, years of education, and time since diagnosis were similar across the whole patient and control groups (*p*-values > 0.05). Comparing across patient groups, svPPA patients tended to be younger (median [IQR] age in years = 65 [63–70]) than both the lvPPA (73 [67–79], *p* = 0.01) and nfvPPA patients (71 [66–73], *p* = 0.09). Symptom duration was longer for svPPA (mean [SD]: 5.8 [4]) than lvPPA (2.4 [2], *p* = 0.009), but not nfvPPA (3.1 [2], *p* = 0.409). Cognitive characteristics revealed by BDAE and ACE scores per PPA subtype are presented in Table 2.

**Table 2.** Demographics and general cognitive characteristics for each PPA subtype and healthy controls.

|  | **lvPPA** | **nfvPPA** | **svPPA** | **Controls** |
| --- | --- | --- | --- | --- |
| No. of participants | 21 | 17 | 16 | 30 |
| Age, Mean [SD] | 73 [67-79] | 71 [66-73] | 65 [63-70] | 68 [65-70] |
| Sex, Male: Female | 15:6 | 6:11 | 8:8 | 18:12 |
| Handedness, Right: Left | 19:1 | 15:2 | 17:0 | 27:3 |
| Education (years), Mean [SD] | 19 [3] | 17 [2] | 19 [2] | 21 [3] |
| Time since diagnosis (years), Mean [SD] | 1.2 [1] | 2 [1.7] | 2.4 [2] | - |
| Language symptom onset (years), Mean [SD] | 2.4 [2] | 3.1 [2] | 5.8 [4] | - |
| **BDAE sub scores, mean [SD]** |  |  |  |  |
| Repetition of single words (/5) | 4 [0.6] | 4 [1] | 4 [0.8] | 5 [0] |
| Auditory comprehension (/16) | 15 [2] | 14 [4] | 11 [3] | 16 [0.2] |
| Picture-word matching (/4) | 3 [1] | 3 [1] | 2 [1] | 4 [0.3] |
| Repetition of sentences (/2) | 1 [0.6] | 1 [0.7] | 2 [0.6] | 2 [0] |
| Boston Naming Test (/15) | 8 [4] | 9 [5] | 3 [3] | 14 [0.4] |
| Oral Reading (/15) | 14 [2] | 12 [5] | 13 [3] | 15 [0] |
| **ACE-III/R sub scores mean [SD]** |  |  |  |  |
| Attention (/18) | 12 [3] | 13 [5] | 15 [2] | 18 [0.6] |
| Memory (/26) | 9 [7] | 14 [8] | 9 [5] | 25 [0.7] |
| Fluency (/14) | 4 [3] | 3 [3] | 4 [2] | 13 [0.3] |
| Language (/26) | 18 [5] | 18 [6] | 11 [3] | 26 [0.4] |
| Visuospatial (/16) | 12 [2] | 12 [5] | 15 [1] | 16 [0] |

lvPPA; Logopenic variant PPA, nfvPPA; non-fluent variant PPA, svPPA; semantic variant PPA; BDAE, Boston diagnostic aphasia examination; ACE, Addenbrooke's cognitive examination; SD, standard deviation.

### Test characteristics

Administration of the MLSE took an average [SD] of 19 [3] minutes, with lvPPA taking the longest at 20 [3] minutes, followed by svPPA at 19 [2] minutes, and nfvPPA 18 [2] minutes.

A two-way mixed effects model (people effects are random and measures effects are fixed) showed scoring decisions made by the three independent raters to be highly consistent, with an ICC index of 0.95 (*p* < 0.0001).

The reliability of the MLSE against the BDAE and ACE for all participants resulted in a Cronbach’s Alpha score of 0.908. Convergent validity produced correlations ranging from *r* = 0.603 – 0.669. Correlation coefficients for each test pair were: repetition of single words and the repetition component of the repeat and point task (*r* = 0.665); auditory comprehension vs. the pointing component of the repeat and point task (*r* = 0.669); repetition of sentences and the sentence repetition task (*r* = 0.613); Boston naming test vs. the naming task (*r* = 0.663); oral reading vs. the reading task (*r* = 0.603); and working memory components of the ACE-III/R with the sentence repetition task (*r* = 0.632). The *p-*values were < 0.001 for all correlations.

### Language profiles

Scores grouped by diagnosis in each of the five linguistic domains are presented in Figure 2 along with group medians and IQRs for individual domains and overall MLSE score. The average total MLSE scores (median [IQR]) were as follows: svPPA patients = 79 [76–82]), lvPPA = 78 [71–84] and nfvPPA 67 [55–76]: F(3,80) = 137.11 (*p* < 0.001). These overall scores were higher in svPPA and lvPPA compared to nfvPPA (*p* = 0.002 and *p* = 0.019 respectively). The distribution of individual domain scores (converted to percentage of maximum score) by PPA syndrome are shown in Figure 3. There were significant group differences associated with all five linguistic domains:

**Figure 2.**
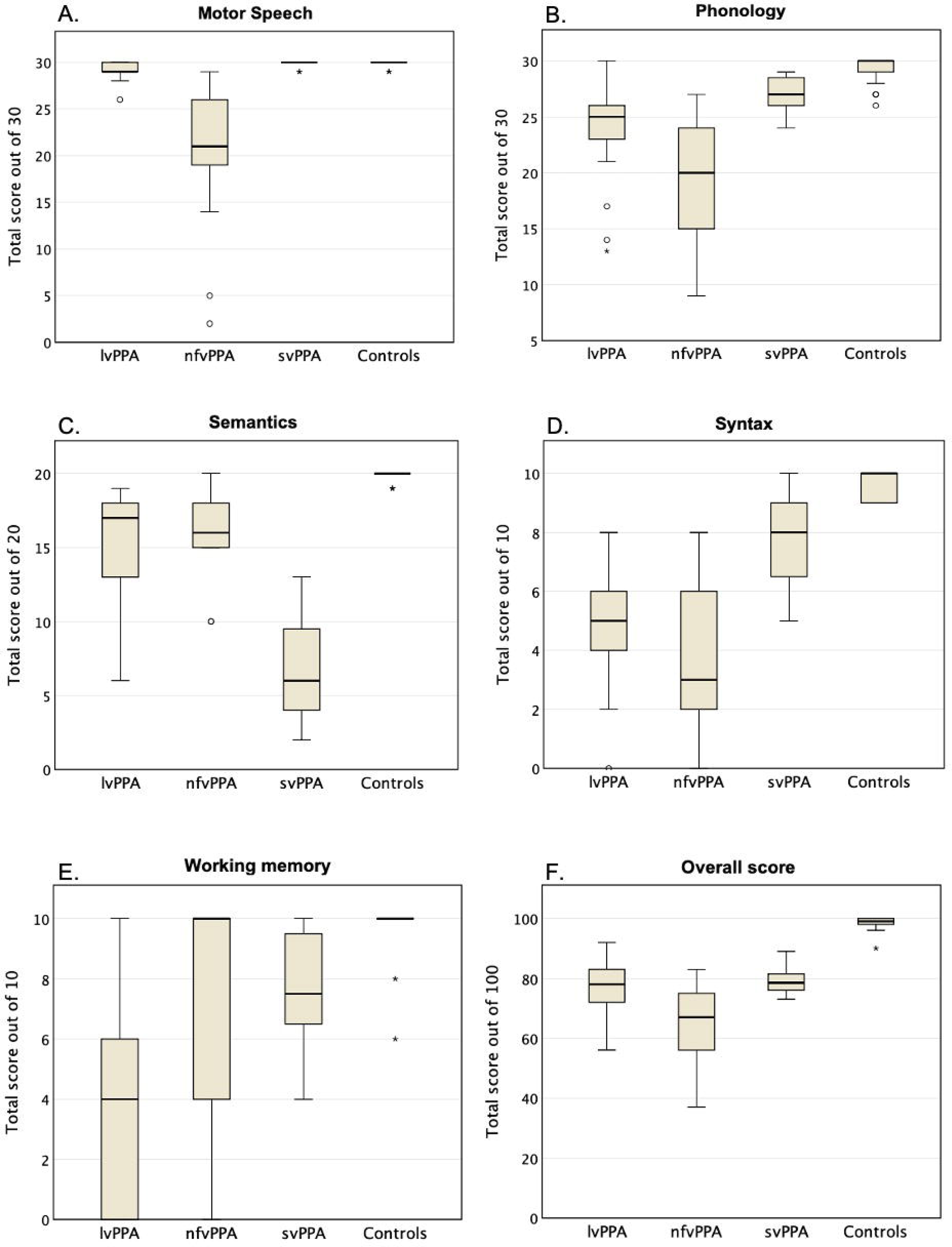
MLSE domain results by diagnosis. The o symbol denotes outliers and * denotes extreme outliers within the groups.

**Figure 3.**
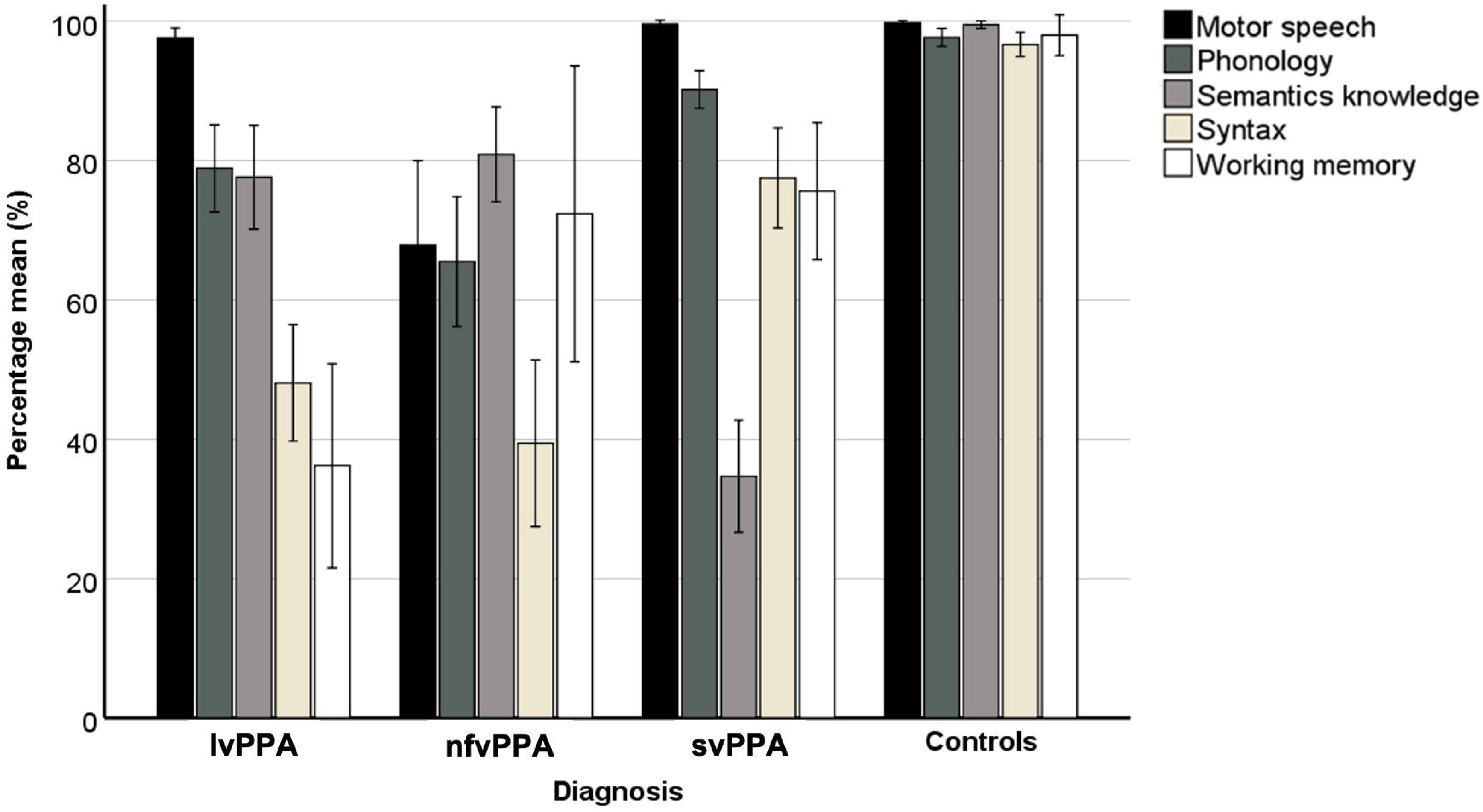
MLSE results illustrating the outcome of the five linguistic domains grouped by PPA subtype and healthy controls. lvPPA; logopenic variant PPA, nfvPPA; non-fluent variant PPA, svPPA; semantic variant PPA.

- <u>Motor speech</u> F(3,80) = 11.72 (*p* < 0.001): the nfvPPA group (percentage mean [SD], 67 [25]) scored significantly lower than both lvPPA (97 [3]) and svPPA (99 [1]), (both *p* < 0.001), and there was a marginal difference in motor speech scores between lvPPA and svPPA (*p* = 0.066).
- <u>Phonology</u> F(3,80) = 30.83 (*p* < 0.001): the nfvPPA group (65 [19]) scored lower than lvPPA (78 [14]) but this was not statistically significant (*p* > 0.05). However, both the nfvPPA group and the lvPPA group scored significantly lower than svPPA (90 [5]) (*p* < 0.01 for both contrasts).
- <u>Semantic knowledge</u> F(3,80) = 102.05 (*p* < 0.001): svPPA patients (34 [16]) scored significantly lower than lvPPA (77 [17]) and nfvPPA patients (80 [14]) (*p* < 0.001 for both contrasts). There was no significant difference in semantic knowledge scores between lvPPA and nfvPPA patients (*p* > 0.05).
- <u>Syntax</u> F(3,80) = 74.11 (*p* < 0.001): scores were significantly lower in lvPPA patients (48 [19]) and nfvPPA patients (39 [24]) than in the svPPA group (76 [14]), (both *p* < 0.001). There was no significant difference in syntax domain scores between nfvPPA and lvPPA patients (*p* > 0.05).
- <u>Working memory</u> F(3,80) = 28.06 (*p* < 0.001): scores were lowest in the lvPPA group (36 [33]) and statistically different from both nfvPPA (72 [43]) and svPPA (75 [19]), (*p* < 0.05 and < 0.001 respectively). There was no significant difference in working memory domain scores between nfvPPA and svPPA (*p* > 0.05).

### Diagnostic accuracy

ROC analysis (see Figures 4a – 4c) revealed that phonology (area under curve (AUC) = 0.77), syntax (AUC = 0.84) and working memory (AUC = 0.89) were the best parameters for the diagnosis of lvPPA (all *p*-values < 0.001). For the diagnosis of nfvPPA, motor speech (AUC = 0.99), phonology (AUC = 0.90) and syntax (AUC = 0.88) were all good parameters (all *p-* values < 0.001), while semantic knowledge (AUC = 0.99) was the best parameter for the diagnosis of svPPA (*p* < 0.001).

**Figure 4.**
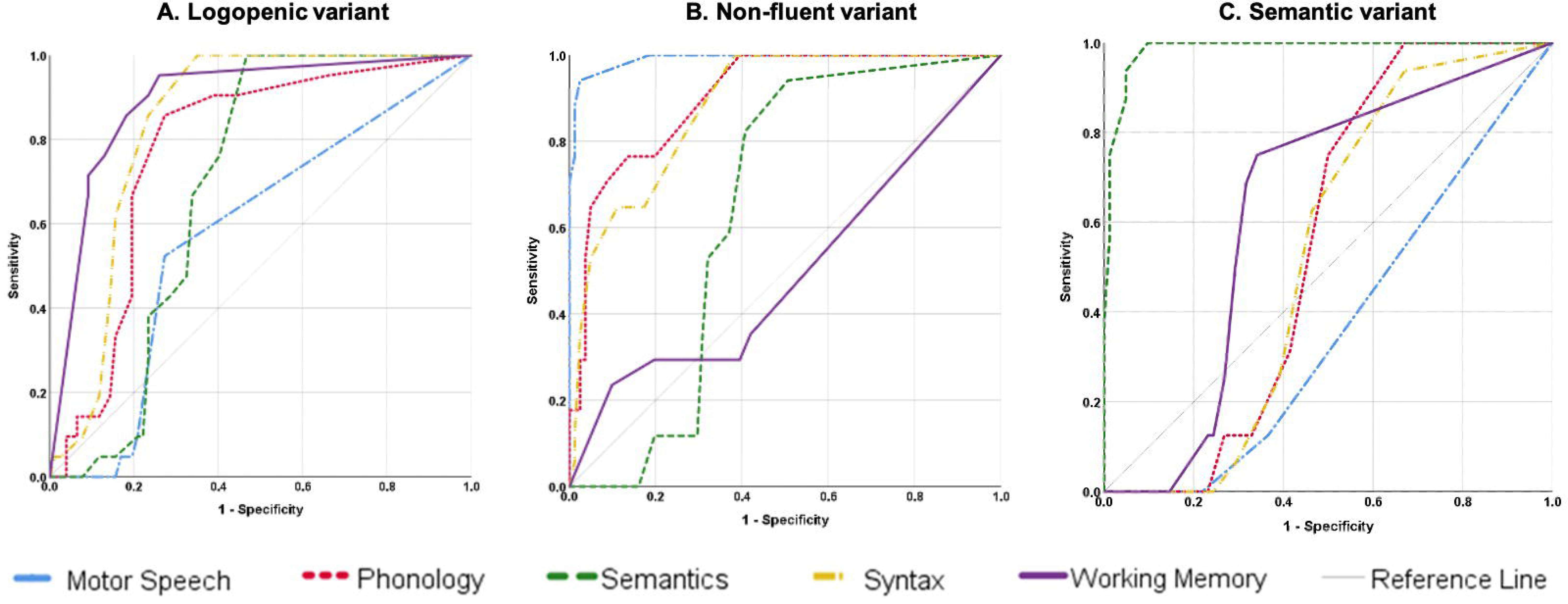
Independent ROC curves demonstrating the accuracy of all five linguistic domain for each PPA subtype.

### Machine learning classification

To further explore the diagnostic accuracy of the MLSE, a robust machine learning method for feature selection and random forest tuning was conducted, based on the five linguistic domains. The predictive capacity of the resulting model was excellent, with an overall accuracy of 0.96. All controls were correctly classified and there were no false positives. Diagnostic accuracies for each of the three syndromes were: 0.92 for lvPPA (89% correctly classified; 1 patient misclassified as nfvPPA; 1 false positive from the svPPA group); 0.93 for svPPA (86% correctly classified; 1 patient misclassified as lvPPA); 0.98 for nvfPPA (100% correct classification, and one false positive from the lvPPA group), Table 3.

**Table 3.** Confusion matrix for predicting PPA diagnosis for 34 participants using Random Forests classification. The overall balanced accuracy of the model was 0.958.

|  |  | *Predicted diagnosis* |  |  |  |  |
| --- | --- | --- | --- | --- | --- | --- |
|  |  | lvPPA, n (%) | nfvPPA, n (%) | svPPA, n (%) | Controls, n (%) | Accuracy |
| *Actual diagnosis* | lvPPA, n (%) | **8 (89)** | 1 (11) | 0 (0) | 0 (0) | 0.924 |
|  | nfvPPA, n (%) | 0 (0) | **7 (100)** | 0 (0) | 0 (0) | 0.981 |
|  | svPPA, n (%) | 1 (14) | 0 | **6 (86)** | 0 (0) | 0.928 |
|  | Controls, n (%) | 0 (0) | 0 (0) | 0 (0) | **11 (100)** | 1.000 |

lvPPA; Logopenic variant PPA, nfvPPA; non-fluent variant PPA, svPPA; semantic variant PPA

A final set of feature rankings for each domain was selected from the results of the training (k-fold) procedure and used in the evaluation of the unseen, out-of-sample set., supplemental material B. Balanced accuracy (the arithmetic mean of sensitivity and specificity) varied as the number of domains reduced. The svPPA and control models showed highest balanced accuracy when using all five domains. The nfvPPA model showed highest balanced accuracy when using four domains (motor speech, phonology, syntax and working memory: 0.943). The lvPPA model achieved highest balanced accuracy with three domains (syntax, working memory and motor speech: 0.944). (See supplemental material B for a detailed breakdown of these figures).

While the random forest classifier was robust and accurate, it does not produce readily interpretable diagnostic rules. A decision tree structure was therefore selected from among the random forests as a guide to manual classification of PPA subtypes from unseen testing data,Figure 5. The tree was selected in view of its accuracy, simplicity and the fact that a diagnosis was made from all five linguistic domains. The decision tree correctly classified 91% (31/34) of the patients and controls whose data were not included in the training set. Two misclassifications were in the lvPPA group (lvPPA2 and lvPPA20 misclassified as nfvPPA). Both of these lvPPA patients scored highly in the working memory and phonology domains. One svPPA patient was misclassified as lvPPA. This svPPA patient showed deficits in the syntax and working memory domains.

**Figure 5.**
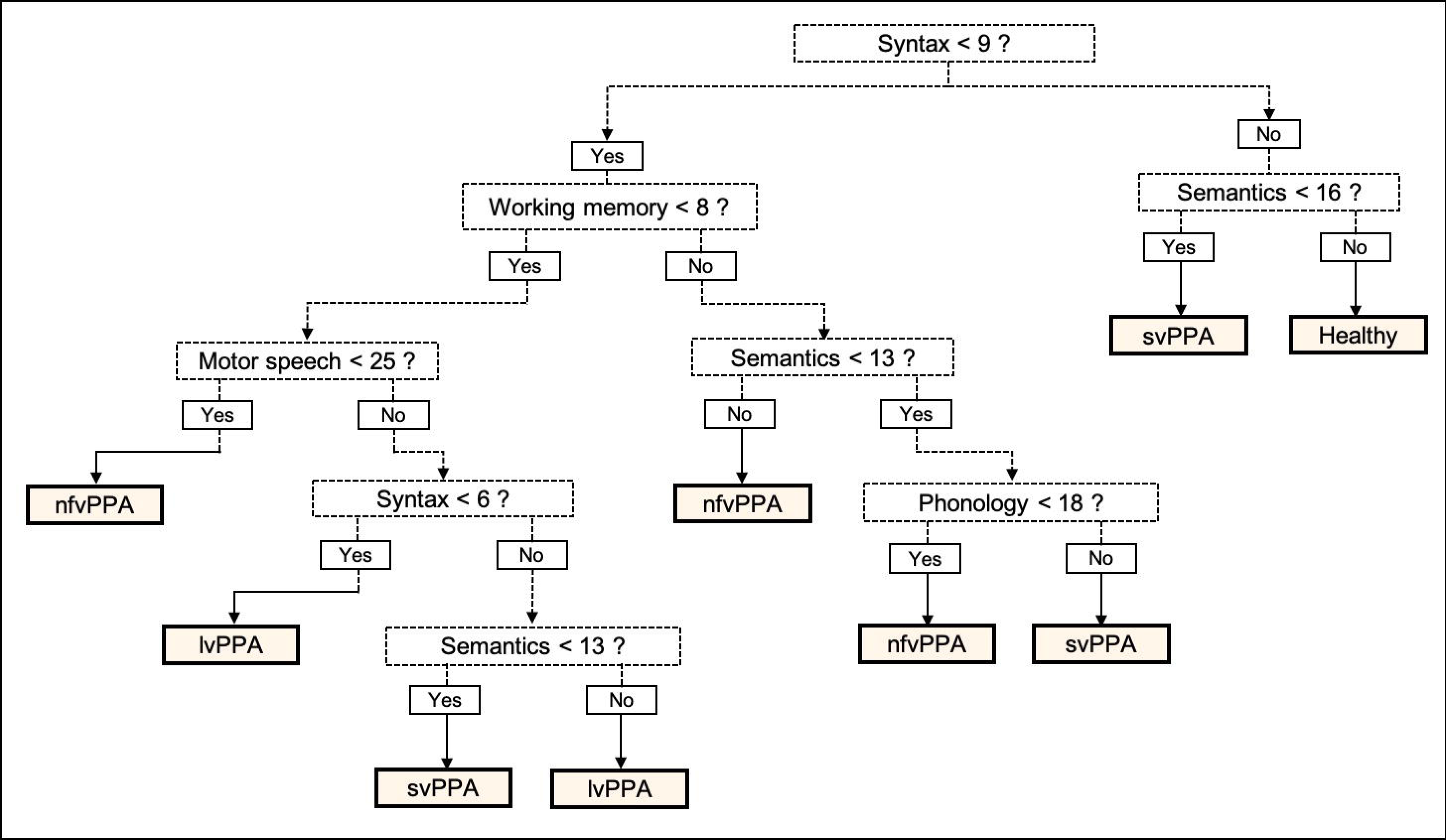
Decision tree using the scores of the five linguistic domains to classify PPA subtypes from the out of sample data. This decision yielded correct classification of 91% (31/34 participants) from the out of sample data (9 lvPPA, 7 svPPA, 7, nfvPPA, 11 controls). lvPPA; logopenic variant PPA, nfvPPA; non-fluent variant PPA, svPPA; semantic variant PPA.

## Discussion

This paper reports the motivation, theoretical assumptions, structure, and diagnostic properties of a novel clinical instrument designed to aid the screening, diagnosis and classification of patients with primary progressive aphasia. The MLSE was motivated by the need for a measure of language competences in PPA that is short, reliable and reproducible, and differentially sensitive to three well-described clinical subtypes of PPA. In its current form, the test enables a clinician efficiently to make quantitative assessments of components of linguistic competence that underpin the diagnoses of the semantic, non-fluent and logopenic variants.

The basic components of linguistic competence – articulation, phonology, semantics, syntax and auditory verbal working memory – are differentially impaired across clinical variants of the syndrome (Hodges and Patterson, 1996; Vonk *et al*., 2019; Lukic *et al*., 2019; Henry *et al*., 2018; Clark *et al*., 2005; Grossman, 2012; Charles *et al*., 2014). The MLSE comprises 11 simple linguistic assays, from which a patient’s profile can be constructed using errors in each of these domains. This approach to scoring amplifies and diversifies the clinical information available from any single test condition, while remaining brief (taking an average of 19 minutes) to administer.

While the assignment of error types is, in principle, subject to disagreements between individual assessors, the development of simple, rule-based guidance led to a high degree of reliability and consistency among raters. Moreover, to ensure the validity of error-based measurement, established assessment instruments (the BDAE and ACE-III) were administered in parallel, and measures relevant to all five domains correlated with the performance levels quantified using the error-based method.

Using the error-based scoring approach, the MLSE was able to distinguish patients with mild PPA from age-matched, control participants with 100% accuracy. Using the different distributions of error-types across the three PPA variants represented in the patient cohort, the MLSE was able to classify 21 of the 23 patients (91%) from the out-of-sample group to their correct diagnosis.

Random forest classification provides a robust statistical method to demonstrate classification accuracy, though it does not provide easily applicable diagnostic rules. As an aid to clinicians, therefore, an additional step for selecting a decision tree was implemented to identify a robust but simple decision structure for manual clinical diagnosis in this cohort. Clinical decision analysis using decision trees became active in the 1990’s, and various methodologies –- such as deep learning and random forest –- have been developed and are being applied to healthcare medical treatment (Breimann, 2001). Although the decision tree presented here provides a further example of the transferability of machine learning models for clinical assistance, we also acknowledge that optimal accuracy was not achieved on the basis of the available data. The models accuracy could therefore be improved by adding data, but an alternative approach would be for the full model to be made available in script format so that the most accurate output could be calculated rapidly on a computer or smart phone for any new combination of domain scores. We intend to add this functionality to the test website http://www.mlsexam.com in the near future.

For an experienced clinician, svPPA can be a relatively straightforward diagnosis, and the group’s performance on the MLSE reproduced the characteristic and relatively isolated impairment of semantic knowledge on which this diagnosis is largely based. More challenging has been the distinction between nfvPPA and lvPPA (Sajjadi *et al*., 2012), as phonology is impaired in both syndromes. That the MLSE can distinguish between these two syndromes is largely due to the fact that motor speech and working memory performance are also quantified, and contribute to a 0.98 accurate classification of nfvPPA, with only one lvPPA placed erroneously into this group.

MLSE performance may substantially aid early and accurate diagnosis. An important contribution to clinical practice is that the MLSE provides a consistent descriptive vocabulary with which clinicians can characterise patients with language disorder of any aetiology (Ingram *et al*., 2020). Whilst the patients reported here were included in an MLSE validation cohort because their language disorder was clearly consistent with either the semantic, the nonfluent, or the logopenic variant of PPA, it is well known that some presentations are not easily assigned to any of these categories (‘mixed PPA’) (Grossman, 2010; Vandenberghe, 2016).

The MLSE may also aid the clinical assessment of other conditions in which compromised language accompanies movement disorders (Peterson *et al*., 2019), dementia (Ahmed *et al*., 2012) or behavioural change (Ash *et al*., 2006; Hardy *et al*., 2016). A well-documented phenomenon is for a patient to present with ‘classic’ nfvPPA and later develop the motor features of corticobasal syndrome (CBS) (Santos-Santos *et al*., 2016) and mixed syndromes are common (Murley *et al*., 2020). A related prodromal phase has been described for progressive supranuclear palsy (PSP) (Whitwell *et al*., 2019; Josephs *et al*., 2005; Josephs and Duffy, 2008; Peterson *et al*., 2019). The development of frontal features of disinhibition and/or obsessionality following presentation with ‘pure’ svPPA is also a common clinical sequence (Ding *et al*., 2020; Murley *et al*., 2020).

Two patients from the current cohort illustrate the fact that the overlap between PPA and AD is more complex than the well-known association between lvPPA and AD pathology (Ahmed *et al*., 2012). Prominent anomia, fluent but empty speech, and impaired semantic knowledge led to a clinical diagnosis of svPPA in patients svPPA2 and svPPA3, yet their performance on the MLSE showed that these features were combined with a low working memory score that was atypical for the group. Biomarkers of AD pathology were later identified in the CSF of both these patients.

We have shown how a machine learning algorithm can learn the relationship between data points of the five linguistic domains and that the features on which this learning was based coincided with *a priori* definitions of the syndromes (Gorno-Tempini *et al*., 2011; Lukic *et al*., 2019; Henry *et al*., 2018). An advantage of the random forest classifier lies in the assessment of data containing irregular samples or missing data points. It can outperform support vector machines and linear mixed effects methods and is thus an effective choice for this type of classification challenge (Moore *et al*., 2019). We summarised the output of the random forest method by representing the rules learned after training in a decision-tree format, while noting that the outputs of the model would be improved with a larger sample size.

Further data collection and analyses are already in progress to determine: i) whether the MLSE can be incorporated into ‘real-world’ clinical or neuropsychological consultations with equivalent degrees of accuracy by non-specialist assessors working with error definitions that, particularly in the case of motor speech, are relatively unsophisticated; ii) whether and to what extent a patient’s profile and/or total score on the MLSE are sensitive to progression of the degenerative process; iii) whether the patterns of domain competence show the expected spatial correlations with regional grey-matter atrophy on MR imaging; and iv) whether the MLSE can be adapted for use in other languages taking account of differential item familiarity, language-dependent vulnerability of different linguistic domains (Canu *et al*., 2020), and the nature of the correspondence between written representations and phonological forms (Hodges *et al*., 1992; Woollams *et al*., 2007; Fushimi *et al*., 2009).

Versions of the MLSE for Italian- and Spanish-speaking populations have been developed and comparisons of the performance of the instrument across these languages are in preparation. We encourage the development of versions in other languages, including those outside the Indo-European family. In the meantime, the test methodology is available and free for non-commercial research under a Creative Commons Licence.

## Conclusions

The MLSE is a new, short (20 minutes) cognitive assessment for the screening and differential diagnosis of primary progressive aphasia, with an innovative scoring system which maximises the clinical information available from a single test. This fast and reliable tool requires minimum clinical expertise to administer. The MLSE can support diagnosis of the three archetypal syndromes of PPA and provide a detailed breakdown of the linguistic profile for mixed PPA cases. The test as described here is for British English, but versions are already available in Italian, Castilian-Spanish, Argentinian-Spanish, American-English and Farsi, with more languages to come. This brief clinical instrument also holds promise for language assessment in other neurological disorders that can affect language, including movement disorders, Alzheimer’s diseases and stroke.

## Data Availability

Anonymised data are available on reasonable request for academic purposes.

## Acknowledgements

We thank all of the patients, their families and carers for their support in this research study. We also thank the Join Dementia Research team for facilitating recruitment for healthy volunteers.

## Supplementary material

Supplemental material A: MLSE Administration and Scoring manual.

Supplemental material B: Feature selection machine learning model of linguistic domains using random forest classification.

